# Healthcare Provider Perceived Competency in Postpartum Hemorrhage Management: Insights from a Quantitative Survey in Uganda’s National Referral Facility

**DOI:** 10.1101/2025.06.10.25329371

**Authors:** Steven Baguma, Kagawa N. Mike, Susan Obore

**Affiliations:** Makerere University College of Health Sciences; Mulago Specialized Women and Neonatal Hospital

**Keywords:** Knowledge, Confidence, Managing PPH, Healthcare providers

## Abstract

**Background:** Postpartum hemorrhage (PPH) remained a significant global health challenge, affecting over 14 million women annually and causing approximately 70,000 maternal deaths worldwide. This study assessed the perceived competency of healthcare providers in managing postpartum hemorrhage at Kawempe National Referral Hospital.

**Methods:** A facility-based cross-sectional study was conducted at Kawempe National Referral Hospital using quantitative data collection methods. The study population comprised 221 obstetric healthcare providers, including 96 midwives, 20 nurses, 18 medical intern doctors, 73 residents in obstetrics and gynecology, and 14 specialists. Data were collected using a pretested structured interviewer-administered questionnaire, and healthcare practices were assessed using the E-MOTIVE bundle. Data analysis was performed using STATA version 18. Logistic regression analysis was conducted to test associations between dependent and independent variables, with a P-value <0.05 considered statistically significant.

**Results:** The study revealed that 81% of healthcare providers demonstrated good knowledge of PPH management, and 61% were confident in their ability to manage PPH. However, only 39.8% of the participants were deemed competent in PPH management. The study found a positive association between specialized training and competence, with HCWs who had received multiple PPH-related training sessions more likely to be competent. Factors such as being a specialist and having received focused training on active management of the third stage of labor were significantly associated with higher competence levels.

**Conclusion:** The study concluded that while knowledge and confidence in managing PPH were generally high among healthcare providers at Kawempe National Referral Hospital, there was a gap in practical competence. Specialized training in PPH management was a significant predictor of competence, underscoring the importance of targeted, hands-on education. The findings suggest that improving training opportunities and resources, particularly in rural and underserved areas, could enhance the overall competence of healthcare providers in managing PPH, ultimately improving maternal health outcomes.

## Introduction

Postpartum hemorrhage (PPH) remains a significant global health challenge, particularly in low-resource settings, where access to skilled obstetric care may be limited (Likis et al., 2015). Each year, about 14 million women experience PPH resulting in about 70,000 maternal deaths globally (WHO, 2023). The global prevalence of PPH is 6 % and the highest burden is experienced in low-income countries (Ononge, Mirembe, Wandabwa, & Campbell, 2016). The magnitude of PPH in sub-Saharan Africa is high at 10.5 % and causes 25 % of all maternal deaths (Sheldon et al., 2024). In Uganda, PPH is known to affect 9% of all pregnancies and is one of the leading causes of maternal mortality in Uganda (Ononge et al., 2016).

Competence refers to the ability of an individual to effectively perform tasks, fulfill responsibilities, and achieve desired outcomes within a specific context or domain (Campion et al., 2011). It involves the integration of knowledge, skills, abilities, attitudes, and behaviors that enable an individual to perform tasks proficiently and meet established standards or expectations (Campion et al., 2011). In the management of PPH, competent healthcare providers play a pivotal role in preventing severe morbidity and mortality associated with PPH by adhering to best practices, including active management of the third stage of labor, monitoring for early signs of hemorrhage, and initiating appropriate interventions (Althabe et al., 2008).

A knowledgeable health workforce is an essential ingredient in the provision of high-quality care (Rowe, De Savigny, Lanata, & Victora, 2005). According to Rowe et la., health workers ought to have sufficient knowledge of PPH causes, risk factors, signs, and symptoms and evidence-based practices, proficiency in clinical skills such as uterine massage, administration of uterotonic drugs, manual removal of placenta, and surgical interventions and confidence in the management of PPH (Rowe et al., 2005). In the same aspect, healthcare workers’ confidence defined as self-belief and assurance in their ability to perform their roles effectively, make sound clinical decisions, and manage patient care competently, plays a crucial role in healthcare delivery as it influences healthcare workers’ behaviors, attitudes, and interactions with patients, colleagues, and other stakeholders (Angelina, Kibusi, & Mwampagatwa, 2019). Self-confidence and self-belief influences healthcare workers’ decision-making processes, effective communication with patients, families, and colleagues, and is often associated with self-efficacy, components which are all crucial for successful management of PPH patients (Bulndi et al., 2017).

In Uganda and other SSA countries, studies have highlighted variations in healthcare workers’ knowledge of PPH risk factors, signs, and management strategies. While some healthcare providers demonstrate a strong understanding of evidence-based practices for PPH prevention and treatment, others may lack comprehensive knowledge, particularly in rural or underserved areas where access to continuing education and training opportunities may be limited (Namagembe, 2023). However, limited research has been conducted to assess the healthcare provider competency levels in the management of postpartum hemorrhage in Uganda, creating barriers to competency development and maintenance in Ugandan obstetric health care providers. This study assessed the perceived competency of healthcare providers in managing postpartum hemorrhage at Kawempe National Referral hospital, Kampala Uganda.

## Methods

### Study design and setting

A facility-based cross-sectional study design employing quantitative methods of data collection was conducted. The study was conducted at Kawempe National Referral Hospital, a government-founded facility located in Kawempe Division, Kampala District, Uganda. It is located 7 km north of Kampala City, about 5 km north of Mulago National Referral Hospital along Bombo Road on Kampala-Gulu Highway. The facility offers maternal and child health services to citizens across the country and also serves as a medical training institution. It registers an average of 23,000 deliveries per year, of which 8,500 are caesarean deliveries. It is manned by 309 obstetric care providers, including 165 midwives, 15 specialists, 90 residents, and 39 intern doctors. According to data extracted from the hospital database between July to September 2023, postpartum hemorrhage accounted for 41% of all maternal deaths at the facility.

### Study population

All obstetric healthcare providers working in the maternity ward, antenatal clinic, postnatal ward, high-dependency unit, and intensive care unit of KNRH who directly interfaced with patients likely to experience obstetric hemorrhage and consented to participate during the study period were Included in the study. While healthcare providers who were newly transferred, and had not yet managed a significant number of postpartum hemorrhage cases, were excluded from the study

### Sample size

Sample size was determined using Krejcie and Morgan (1970) tables for sample size estimation in studies with finite populations. The sample size for each category was determined and summed up. A 20% attrition for each category was added to cater for non-response and missing information.

| **Participants Per Professional Category.** | **Number Of Obs Care Providers** | **Sample Size Per Category (%)** | **Representative Number After Attrition** |
| --- | --- | --- | --- |
| Midwives | 165 | 113 | 136 |
| Intern doctors | 39 | 32 | 38 |
| Residents OBGYN | 90 | 73 | 88 |
| Specialists | 28 | 14 | 17 |
| **Total** | **309** | **232** | **278** |

After data cleaning process, a total of 221 participants had complete data and were assessed in further statistical analysis.

### Sampling procedure

Stratified sampling was utilized to ensure representation across different professional roles. Within each stratum, convenience sampling methods were employed to select participants, ensuring voluntary participation and minimizing selection bias.

### Data collection tools and procedures

Participants were found in different wards of KNRH, where they were assessed for eligibility. Data were collected using a pre-tested structured, both interviewer and self-administered questionnaires using Kobo-Tools. The questionnaire comprised four sections, Sociodemographic characteristics (age, sex, qualifications, working experience, etc.), Knowledge on PPH management assessed through 25 questions on diagnosis, prevention, and management, with correct and incorrect options, Confidence in handling PPH, assessed via a self-assessment survey rated on a three-point Likert scale and PPH management practices, assessed using reflective practice and recorded in relation to the E-MOTIVE bundle.

### Statistical Analysis

Data entry was done using Kobo Collect and analyzed with STATA Version 18. Descriptive statistics were utilized to assess the demographic characteristics of the participants. Categorical variables were summarized by frequencies and percentages, while continuous variables were summarized by mean and standard deviation or median and interquartile range, depending on their distribution. The results were presented in tables, graphs and Pie charts.

A total of twenty-five questions were utilized to assess knowledge of PPH, with one mark awarded for each correct response. The total number of correct responses was summed up, good knowledge of PPH management was attained with mean score of ≥17 (SD ±2) of 25 total knowledge score.

To evaluate confidence in PPH management, 21 questions were posed (12 for nurses and midwives, 21 for doctors), granting 2 marks and 1 mark if an individual was capable of performing a procedure independently or with assistance respectively while those who could not awarded no mark. The total number of correct responses was summed up, mean score of ≥34 (SD ±6) of 44 total Confidence score for JHO/SHO/Specialist and mean score of ≥16 (SD ±3) of 24 total Confidence score for Midwife/Nurse. For the practice of PPH management, 9 questions were included, where respondents who answered “always” received 2 marks, “rarely or Sometimes” received 1 mark and no mark for “never”. The total sum of responses was summed up and a mean score of ≥15 (SD ±1) of 18 total practice score.

For further analysis, PPH knowledge, confidence, and practices were categorized into a binary variable using the mean score of each section as the cutoff point.

To measure association between dependent and independent variables, bivariate and multivariable analysis using logistic regression was done. Variables with a P-value <0.2 at bivariate level were considered for multivariable analysis. At multivariable level, a P-value <0.05 at 95% confidence interval was considered statistically significant.

### Ethical considerations

Permission was obtained from the Department of Obstetrics and Gynecology. Ethical approval was granted by the Institutional Review Board of Makerere University School of Medicine (**MAK-SOMREC-2024-1031).** Administrative clearance was obtained from Kawempe National Referral Hospital. Written informed consent was obtained from all participants. Privacy and confidentiality were maintained by assigning codes to questionnaires.

## Results

### Sociodemographic characteristics of participants

A total of 221 HCWs were participated in the study with a median age of 34 years (IQR 29-38). Majority were females 134 (60.6%), were married 150 (67.9%) and had a bachelor’s degree 108 (48.9%) and frequently managed PPH in their practice 120 (54.3%) **Table 1**

**Table 1:** Sociodemographic characteristics of participants.

| VARIABLE | FREQUENCY<br>(N=221) | PERCENTAGE |
| --- | --- | --- |
| **Age (Years)** |  |  |
| 20-29 | 62 | 28.0 |
| 30-39 | 111 | 50.2 |
| ≥40 | 48 | 21.7 |
| **Gender** |  |  |
| Female | 131 | 59.3 |
| Male | 90 | 40.7 |
| **Marital status** |  |  |
| Divorced | 3 | 1.4 |
| Married | 150 | 67.9 |
| Separated | 1 | 0.5 |
| Single | 67 | 30.3 |
| **Level of education** |  |  |
| Bachelors | 108 | 48.9 |
| Certificate | 19 | 8.6 |
| Diploma | 65 | 29.4 |
| Masters | 29 | 13.1 |
| **Cadre** |  |  |
| Junior House Officer | 18 | 8.1 |
| Midwife | 96 | 43.4 |
| Nurse | 20 | 9.0 |
| Senior House Officer | 73 | 33.0 |
| Specialist OB/GYN | 14 | 6.4 |
| **Experience working at KNRH** |  |  |
| Less than 1 year | 87 | 39.4 |
| 1-2 years | 45 | 20.4 |
| 3-4 years | 50 | 22.6 |
| More than 5 years | 39 | 17.6 |
| **Frequency of managing PPH** |  |  |
| Frequently | 120 | 54.3 |
| Occasionally | 71 | 32.1 |
| Rarely | 30 | 13.6 |

**Figure 1** demonstrates that majority of the participants (n=200) had training in active management of third stage of labor, management of PPH using E-MOTIVE (n=193) and in basic emergency obstetric care (n=154).

**Figure 1.**
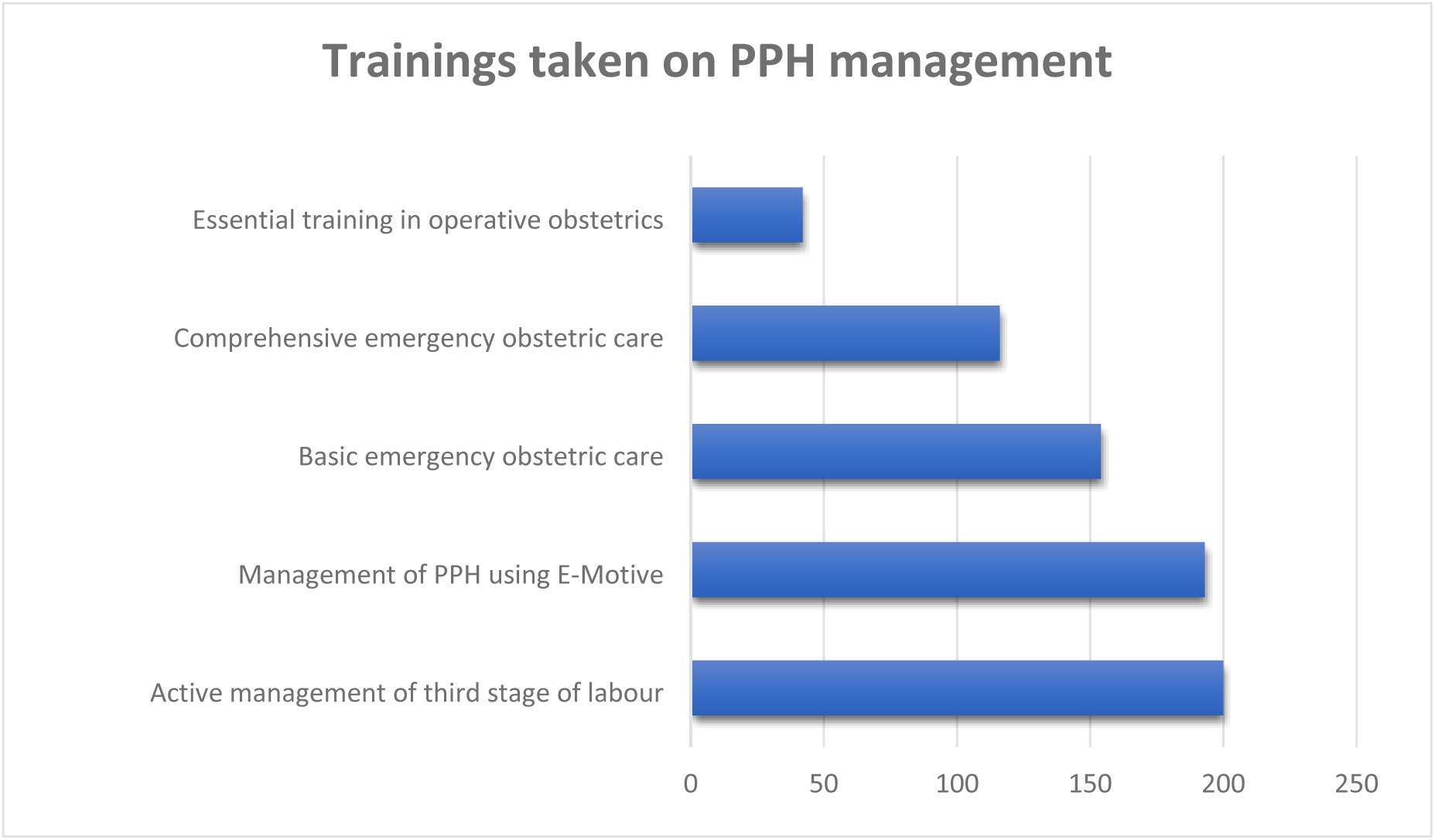
AMSTL-Active management of third stage of labour, BEMOC-Basic emergency obstetric care, CeMOC-Comprehensive emergency obstetric care, ETOO-Essential Training in Operative Obstetrics.

### Knowledge of HCWs on the management of PPH

Three quarters of the participants 180 (81%) had good knowledge of PPH management with mean score of ≥17 (SD ±2) of 25 total knowledge score ***Figure 2***.

**Figure 2.**
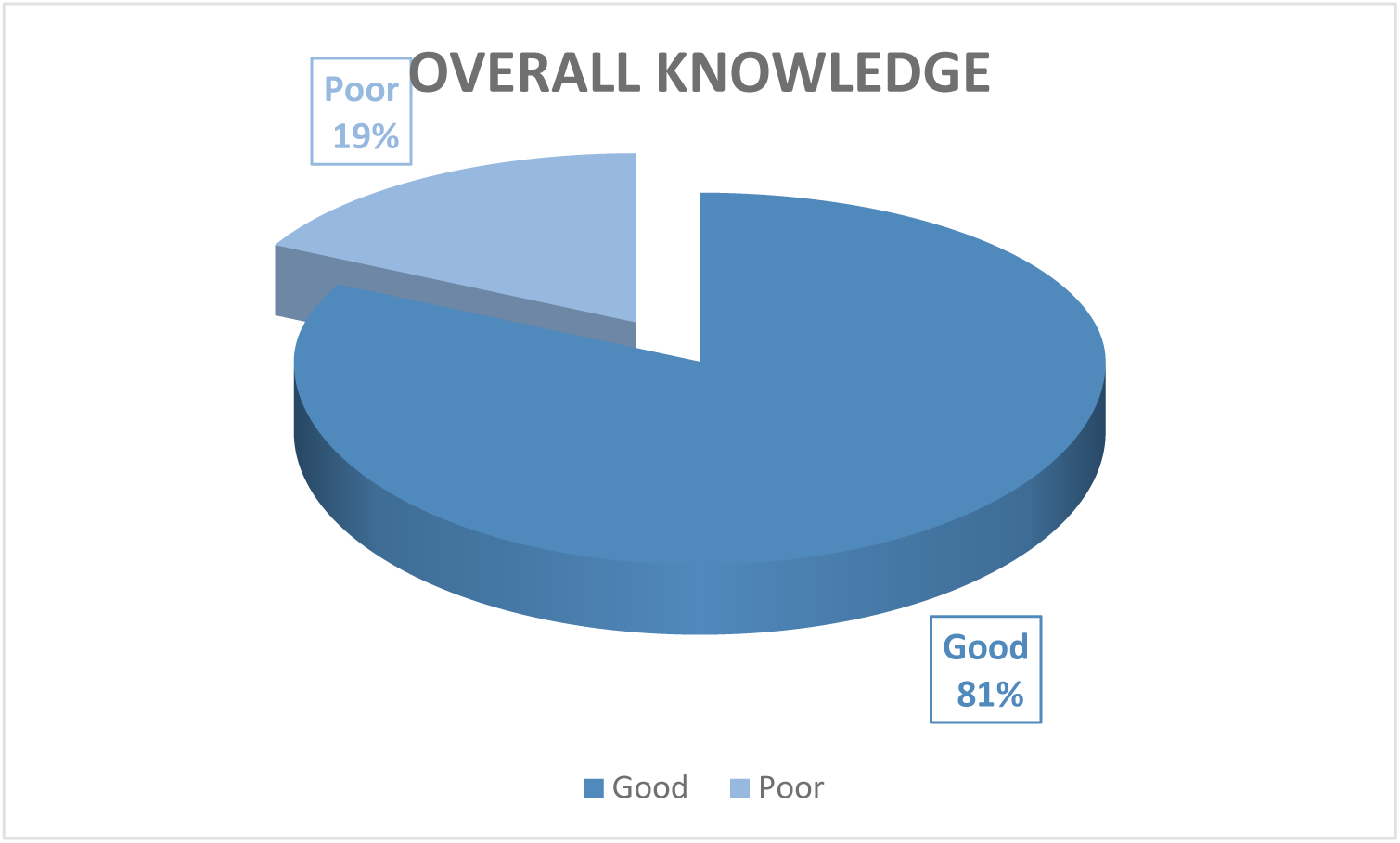
Knowledge of HCWs on the management of PPH.

### HCWs confidence in handling PPH

More than half of participants 134 (61%) were confidence in management of PPH ***Figure 3***.

**Figure 3.**
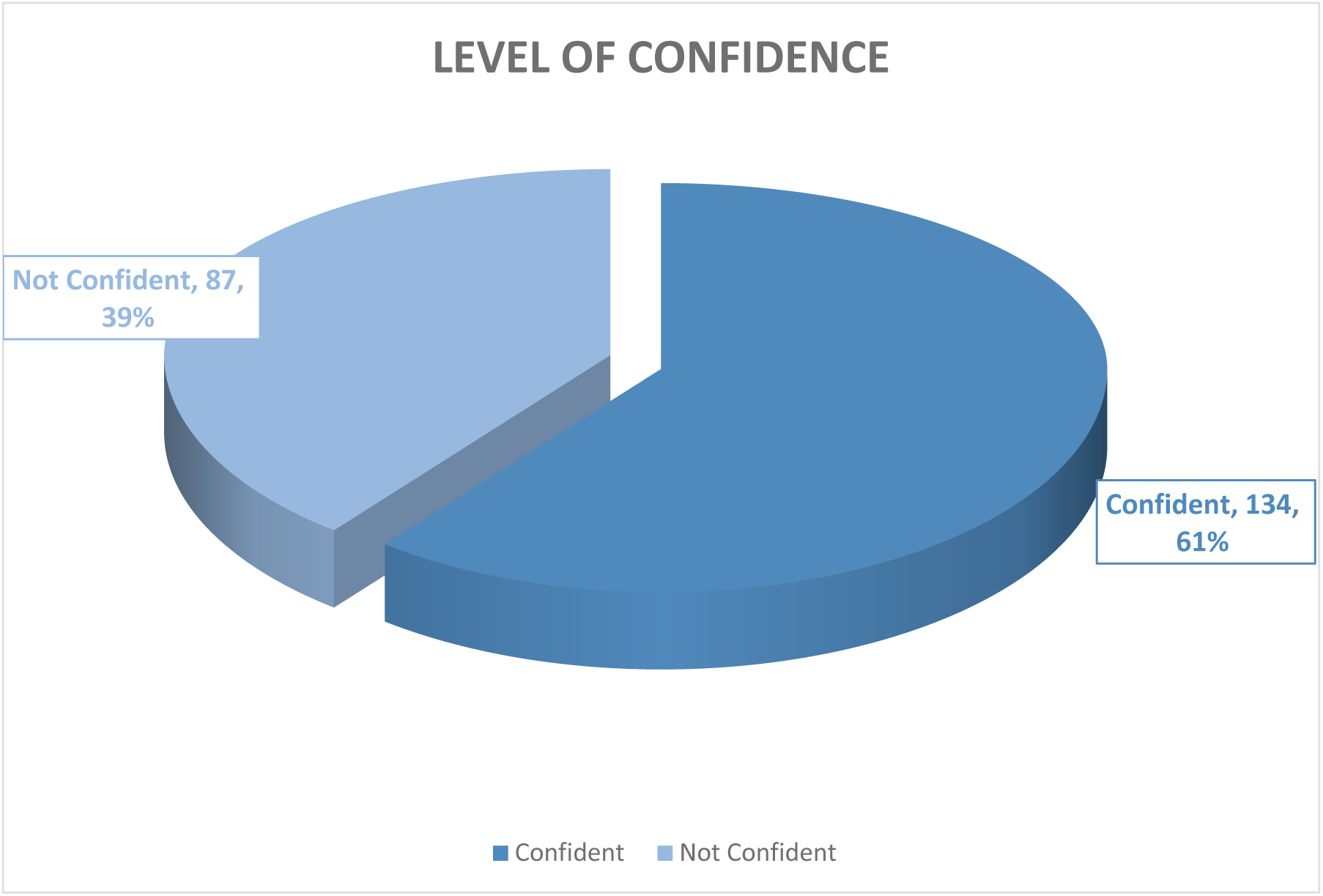
HCWs confidence in handling PPH.

### PPH management practices among HCWs

Three Quarters 159 (71.9%) of HCWs were found to have good practice of PPH management ***Figure 4***.

**Figure 4.**
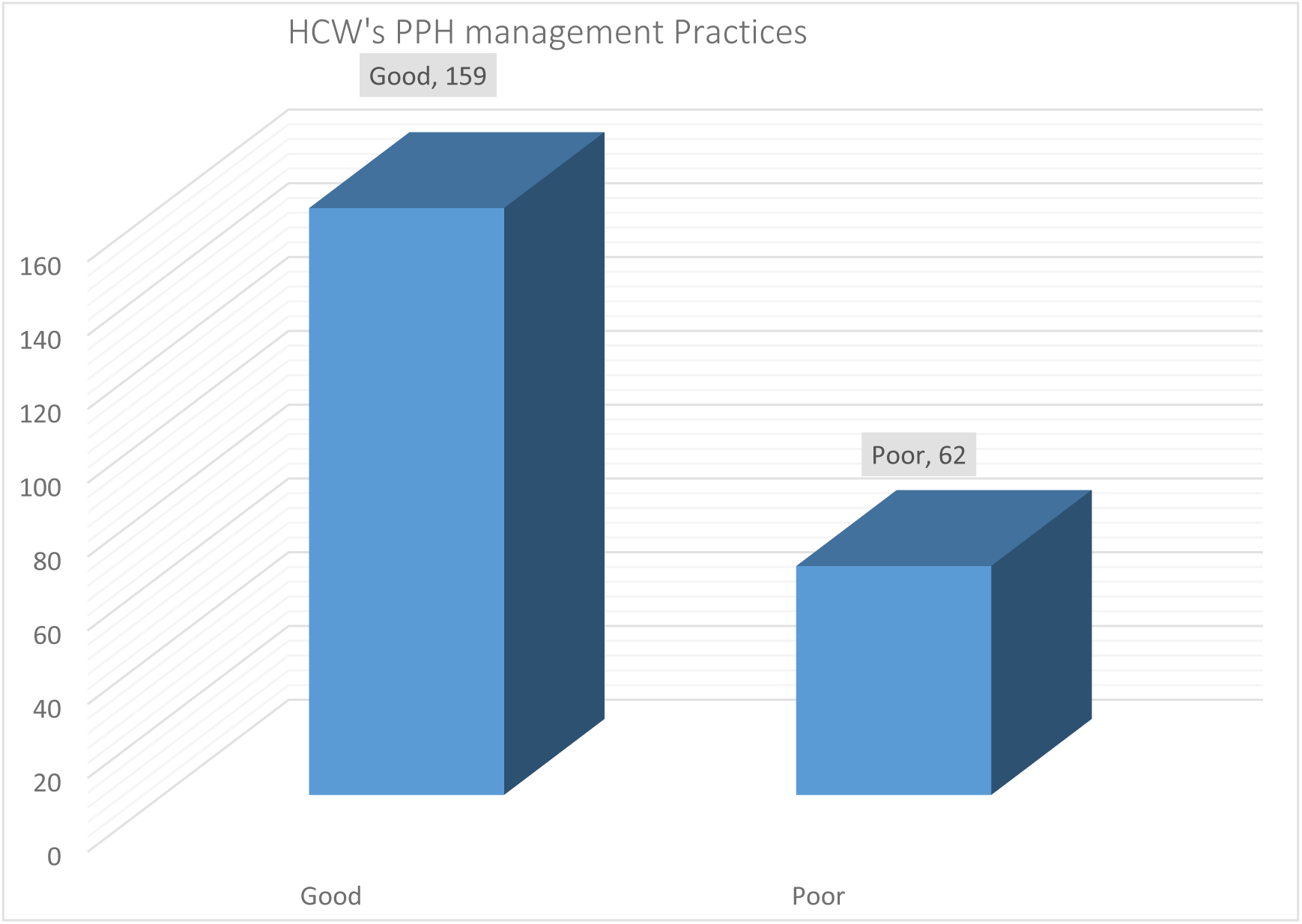
PPH management practices among HCWs.

### HCWs perceived competency in management of postpartum hemorrhage

Of the total 221 respondents, 39.8% were deemed competent, while 60.2% were incompetent **Figure 5**.

**Figure 5.**
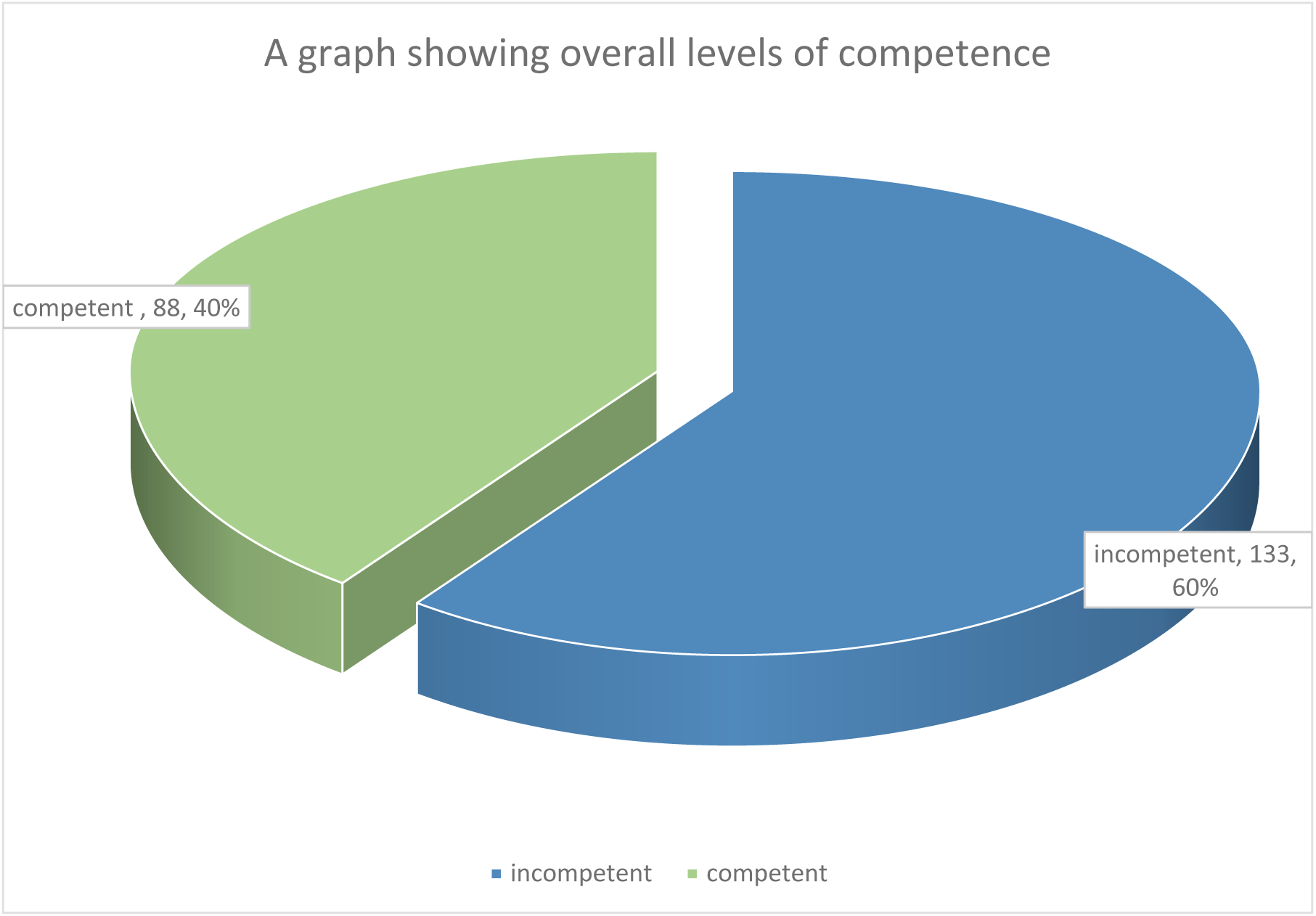
HCWs perceived competency in management of postpartum hemorrhage.

### Competence levels in PPH management among different healthcare cadres

Among the five groups, Junior House Officers (JHOs) and Nurses exhibit the highest levels of incompetency, with approximately 80% classified as incompetent. Midwives also show a majority as incompetent, though to a slightly lesser extent. Senior House Officers (SHOs) display a more balanced distribution, with around 60% deemed incompetent and 40% competent. In contrast, Specialists demonstrate the highest competency levels, with roughly 70% marked as competent and only about 30% as incompetent **figure 6**.

**Figure 6.**
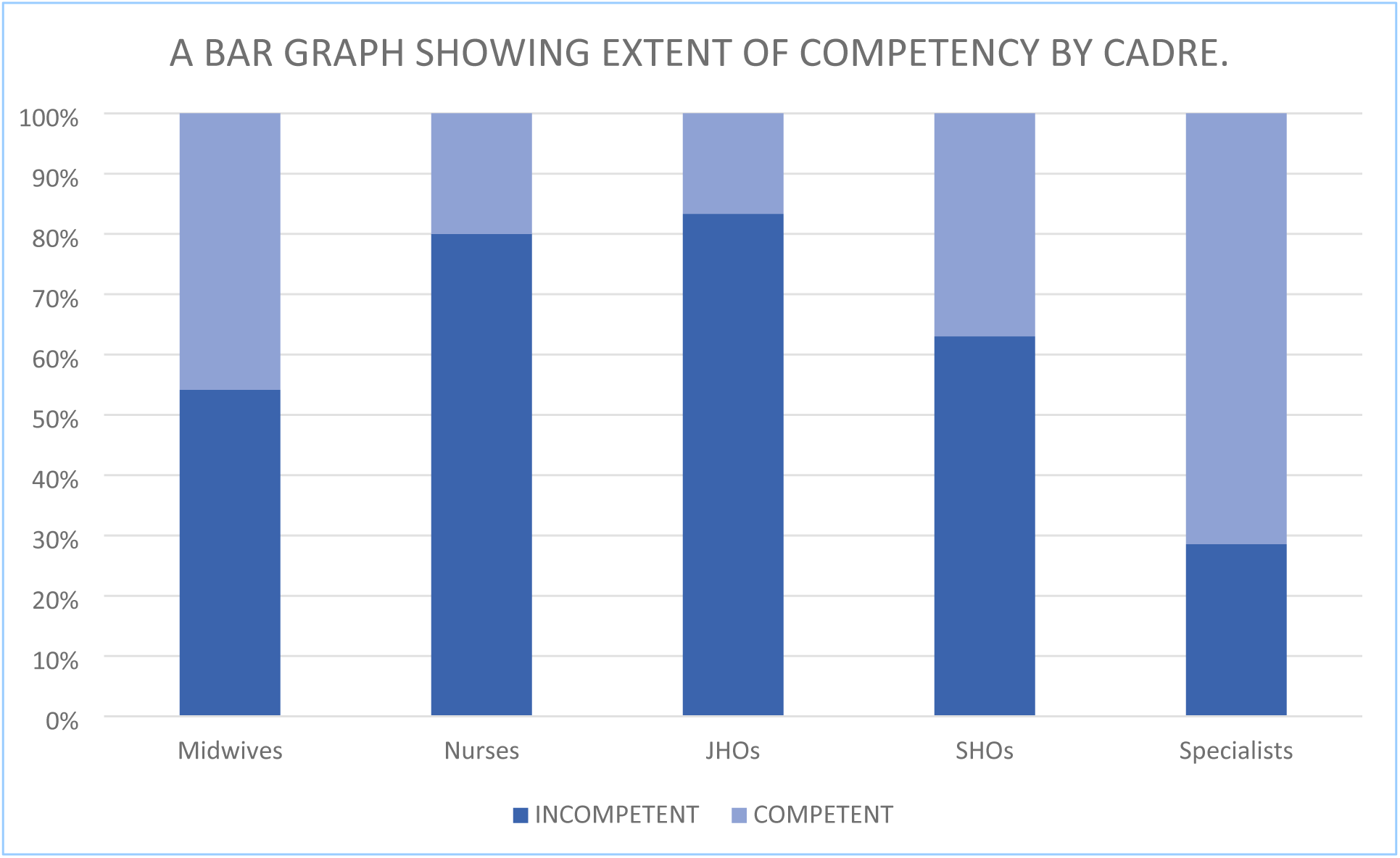
A BAR GRAPH SHOWING EXTENT OF COMPETENCY BY CADRE.

### Factors associated with HCWs perceived competency

Among cadres, midwives comprised the highest proportion of competent staff (50.0%), while nurses and junior house officers (JHOs) had significantly lower odds of competence in the unadjusted analysis (COR = 0.29 and 0.23 respectively), though these associations lost statistical significance after adjustment. For education, those with a master’s degree had the highest competence proportion (20.5%), and while their adjusted odds ratio suggested increased competence, this was not statistically significant (AOR = 3.35, 95% CI: 0.57-19.63, p = 0.180). Notably, health workers who had taken two PPH-related trainings were significantly more likely to be competent than those with only one training (AOR = 0.34, 95% CI: 0.12–0.97, p = 0.044) **Table 2**.

**Table 2:** Bivariate and multivariable analysis of Factors associated with HCWs perceived competency.

| Variable | Level of Competence |  | COR (95%CI) | P-value | AOR (95% CI) | P-value |
| --- | --- | --- | --- | --- | --- | --- |
|  | Competent | Incompetent |  |  |  |  |
|  | N=88<br>(39.8%) | N=133<br>(60.2%) |  |  |  |  |
| Level of Education |  |  |  |  |  |  |
| Certificate | 7 (8.0%) | 12 (9.0%) | Ref | Ref | Ref | Ref |
| Diploma | 31 (35.2%) | 34 (25.6%) | 1.56(0.54-4.47) | 0.405 | 2.03(0.71-5.75) | 0.185 |
| Bachelors | 32 (36.4%) | 76 (57.1%) | 0.72(0.26-2.00) | 0.531 | 1.51(0.39-5.90) | 0.554 |
| Masters | 18 (20.5%) | 11 (8.3%) | 2.80(0.84-9.28) | 0.091 | 3.35(0.57-19.63) | 0.180 |
| Cadre |  |  |  |  |  |  |
| Midwife | 44 (50.0%) | 52 (39.1%) | Ref | Ref | Ref | Ref |
| Nurse | 4 (4.5%) | 16 (12.0%) | 0.29(0.09-0.94) | 0.041 | 0.37(0.10-1.39) | 0.142 |
| JHO | 3 (3.4%) | 15 (11.3%) | 0.23(0.06-0.87) | 0.030 | 0.22(0.04-1.16) | 0.075 |
| SHO | 27 (30.7%) | 46 (34.6%) | 0.69(0.37-1.29) | 0.249 | 0.52(0.16-1.68) | 0.273 |
| Specialist | 10 (11.4%) | 4 (3.0%) | 2.95(0.87-10.07) | 0.084 | 0.89(0.13-6.39) | 0.909 |
| Training Taken |  |  |  |  |  |  |
| One | 13(14.8%) | 15(11.3%) | Ref | Ref | Ref | Ref |
| Two | 9(10.2%) | 33(24.8%) | 0.31(0.11-0.90) | 0.031 | 0.34(0.12-0.97) | **0.044** |
| Three | 17(19.3%) | 36(27.1) | 0.52(0.21-1.40) | 0.207 | 0.55(0.20-1.48) | 0.237 |
| Four | 49(55.7%) | 49(36.8) | 0.96(0.50-2.68) | 0.740 | 1.19(0.47-3.00) | 0.719 |
| Five | 17(19.3%) | 17(12.8%) | 1.08(0.39-2.97) | 0.886 | 0.95(0.31-2.92) | 0.933 |

## Discussion

A total of 221 healthcare workers (HCWs) participated in this study assessing competence in postpartum hemorrhage (PPH) management. Knowledge levels were high among participants, with 81% demonstrating good knowledge. Competence in PPH management was reported in only 39.8% of respondents. HCWs who had taken two PPH-related trainings were significantly more likely to be competent compared to those with only one (AOR = 0.34, p = 0.044).

The knowledge levels of healthcare providers in this study were notably high, with 81% demonstrating good knowledge on PPH management. This is in line with (Akter et al., 2022) that reported high levels of knowledge among healthcare workers. A study in Kenya revealed that healthcare workers were able to recall 71% of recommended clinical actions for managing PPH. However, it was also noted that deficiencies existed in areas like the management of refractory PPH caused by atonic uterus, a crucial component of PPH management (Henry et al., 2022). The results of this study further align with findings from Sub-Saharan Africa, where a systematic review reported an overall prevalence of healthcare providers’ knowledge on PPH management of 47.975%, with pre-service training, higher degrees, and good practices being significantly associated with better knowledge (Abebe Gelaw et al., 2024). The high knowledge score of the participants in our study could also be attributed to the widespread availability of training programs like the E-MOTIVE protocol for PPH management, AMSTL, BEMOC, CeMOC, and ETOO.

In this study, high levels of confidence observed among participants. This is in line with a study in the United States by (Joseph et al., 2020) which revealed varying confidence levels, with obstetricians and midwives expressing high levels of confidence in routine scenarios. Similarly, this study found that 100% of specialists and 65.6% of midwives were confident in managing PPH, which is comparable to findings from studies in high-resource settings where structured training and clinical experience bolstered providers’ confidence (Parry-Smith et al., 2021). A study by (Khadim et al., 2023) supports the importance of good teamwork and communication in boosting confidence levels and improving clinical outcomes during management of PPH.

While the majority of participants reported good knowledge and confidence, only 39.8% of HCWs demonstrated competence in PPH management. A similar trend was observed in studies from Madagascar and Kenya, where healthcare workers demonstrated good knowledge of some aspects of PPH management but had significant gaps in their overall competency (Bazant et al., 2013; Henry et al., 2022). The discrepancy between knowledge and practice can be attributed to several factors, including limited access to hands-on training, inadequate clinical supervision, and resource constraints, which have been well-documented in low- and middle-income countries (LMICs) (Forbes et al., 2024). This study found that HCWs who attended two PPH-related training sessions were significantly more likely to be competent compared to those who attended only one session. This finding aligns with research indicating that pre-service and in-service training play a critical role in improving knowledge and practices related to PPH management (Abebe Gelaw et al., 2024).

One of the key strengths of this study is its comprehensive and large sample size, involving 221 healthcare workers (HCWs). This enhances the generalizability of the findings and provides a robust representation of the effects of knowledge, confidence, and practices of HCWs in managing postpartum hemorrhage. Despite its strengths, the cross-sectional design limits the ability to infer causality. The reliance on self-reported data, HCWs may overestimate their knowledge and confidence, potentially leading to response bias. The study was conducted at a single institution, which limits the generalizability of the findings to other healthcare settings, particularly in different geographic regions or healthcare systems.

### Conclusions

This study reveals that a significant proportion of HCWs demonstrated good knowledge and high confidence in managing PPH, with specialists and midwives showing particularly strong performance. The study also found that fewer number of training sessions had a negative association with confidence, suggesting that the quality of training and the depth of knowledge may be more influential in boosting HCWs’ confidence than the frequency of training.

### Recommendations

The study underscores the need for comprehensive and specialized training programs focused on all aspects of PPH management, including the recognition of early signs, accurate blood loss estimation, and advanced interventions such as exploratory laparotomy and uterine artery ligation. To improve the practical aspects of PPH management, healthcare institutions must ensure that essential resources are readily available.

## Declarations

### Availability of Data and materials

The datasets used and/or analyzed during the current study available from the corresponding author on reasonable request.

### Ethics approval and consent to participate

Institutional approval was obtained from the Department of Obstetrics and Gynecology. Ethical approval was granted by the Institutional Review Board of Makerere University School of Medicine (MAK-SOMREC-2024-1031). Administrative clearance was obtained from Kawempe National Referral Hospital. Written informed consent was obtained from all participants. Privacy and confidentiality were maintained by assigning codes to questionnaires.

## Data Availability

All relevant data are within the manuscript and its Supporting Information files.

## Acknowledgements.

The team appreciates all participants and research assistants for their invaluable support in this study.

## Author Contributions

Steven Baguma (SB): Principal Investigator. Conceived and designed the study, coordinated data collection, led the analysis and interpretation of findings, and was the lead author of the manuscript.

Kagawa N Mike (KNM): Senior Supervisor. Provided critical input on study design, supervised the research process, contributed to data interpretation, and reviewed and revised the manuscript for important intellectual content.

Susan Obore (SO): Supervisor. Assisted in protocol development, supported field supervision and quality control during data collection, and contributed to manuscript review and editing.

All authors reviewed and approved the final version of the manuscript for submission.

